# A Case Study of Transfer of Lesion-Knowledge

**DOI:** 10.1101/2020.08.19.20178210

**Authors:** Soundarya Krishnan, Rishab Khincha, Lovekesh Vig, Tirtharaj Dash, Ashwin Srinivasan

## Abstract

All organs in the human body are susceptible to cancer, and we now have a growing store of images of lesions in different parts of the body. This, along with the acknowledged ability of neural-network methods to analyse image data, would suggest that accurate models for lesions can now be constructed by a deep neural network. However an important difficulty arises from the lack of annotated images from various parts of the body. Our proposed approach to address the issue of scarce training data for a target organ is to apply a form of *transfer learning*: that is, to adapt a model constructed for one organ to another for which there are minimal or no annotations. After consultation with medical specialists, we note that there are several discriminating visual features between malignant and benign lesions that occur consistently across organs. Therefore, in principle, these features boost the case for transfer learning on lesion images across organs. However, this has never been previously investigated. In this paper, we investigate whether lesion knowledge can be transferred across organs. Specifically, as a case study, we examine the transfer of a lesion model from the brain to lungs and lungs to the brain. We evaluate the efficacy of transfer of a brain-lesion model to the lung, and the transfer of a lung-lesion model to the brain by comparing against a model constructed: (a) without model-transfer (i.e.random weights); and (b) using model-transfer from a lesion-agnostic dataset (ImageNet). In all cases, our lesion models perform substantially better. These results point to the potential utility of transferring lesion-knowledge across organs other than those considered here.

## 1 Introduction

Cancer is one of the deadliest diseases in the world, with tens of millions diagnosed with some form of cancer annually. Early diagnosis is one of the most important factors in its control and prevention. Computer-aided detection (CAD) systems, specifically deep learning models could potentially help radiologists by detecting features that can be missed even by the trained eye.

The performance of deep learning models is often heavily dependent on the data available. Given the extensive expertise required for generating annotated medical data [13], ethical and privacy concerns around sharing it [3], obtaining training data for deep models remains a bottleneck. For cancer, this problem is compounded by the fact that not some organs have sparse data. One approach to deal with the lack of data, is to draw on techniques of *transfer-learning* that exploit a commonality across datasets. If such a commonality exists, then the parts of the model constructed on a larger dataset (the “source” model) could be re-used to construct a model for the smaller dataset (the “target” model). In the case of lesions (tumours), there are clinical reasons to expect some common visual features in tumours across organs: (a) Malignant tumours across the body have irregular boundaries, and benign tumours have clear boundaries and sharp margins; (b) Malignant tumours are often found to have a thickening at the periphery; and (c) Malignant tumours are generally found to have inhomogenous attenuation, and benign tumours are characterised by homogeneous attenuation instead [1]. This suggests that source-models for detecting lesions that are constructed for one organ should be helpful in constructing a target-model for detecting tumours in a different organ. The usual approach to transfer-learning however focuses on the use of large, generic datasets for constructing a source model: it uses a source-model with pre-trained ImageNet weights. However, it has been pointed out that the nature of classification in the ImageNet dataset is far different from medical classification [12]. ImageNet weights are tuned for tasks in which the subject is prominent in the background, tasks such as cancer tumour classification involve local features, such as inhomogeneous intensities and irregular boundaries, and some subtle pattern-based features.

The principal motivation for this paper is provided by the work in [10], where it is shown that transfer of knowledge is possible across diseases. We are also inspired by the paper [8], in which the authors note that if the target task is localisation-sensitive (as it is in our case), the gains of using ImageNet weights for initialisation are limited to only a reduction in convergence time, without a big boost in performance.

In this paper, we study the effect of transfer learning from a different organ versus the standard practice of transfer learning from the ImageNet database. To the best of our knowledge, this kind of study has not been performed for lesion classification. We focus on transfer of lesion-knowledge from brain to lungs, and lungs to the brain. Our results using lesion-specific source data are promising: (1) Performance improves substantially (importantly, recall over the malignant class improves); (2) The time taken for model-construction decreases significantly; (3) Model performance has less variance; and (4) These effects are even more pronounced when target data is limited.

## 2 Note on Transfer Learning

Transfer learning is now a thriving area of application, especially using deep neural networks [14]. In essence, this consists of using a large *source* dataset to identify a deep network structure and weights. This model is then “transferred” to construct a model for *target* data, usually by re-estimating using the target data, the weights associating with higher layers of the network. Since this involves fewer estimates than the entire model, it can normally be done with lesser data than would be needed to estimate all weights in the model. For the purposes of this paper, we take the following high-level view of the transfer learning process:

– We will use “model *m*” to denote the pair (*π, θ*), where *π* denotes the structure of *m* (for example, of a deep network) and *θ* denotes the parameters in *m* (for example, the weights in the network). We will assume structures are drawn from some space *Π* and parameters from a space *Θ*, and *M* = *Π × Θ*. Given an instance **x** from a set of instances *X*, and a model *m*, we assume a function *Predict*: *M* × *X* → *Y*, where *Y* is some set of (class) labels. Then, *Predict*(**x** *m*), is to be read as “the value of *Predict* on an instance **x** given model *m*”.
– Let 𝒟 denote the set of subsets of *X × Y*. We use use the term “model construction” to mean a function *Learn*: 𝒟 → *M*. Thus, *m* = *Learn*(*d*) is to be read as “the model *m* constructed by *Learn*, using data *d*”. Using the function *Transfer*: *𝒟* × *M* → *M*, by “model transfer from source to a target”, we will mean the composition *Transfer ◦ Learn*. That is, given *source* data *d_s_*, and *target* data *d_t_*, a model transfer from source to target is the model *Transfer*(*d_t_ Learn*(*d_s_*)). Both *Learn* and *Transfer* are conditional on the definition of source-specific and target-specific loss functions. We omit this detail here.
– Testing model performance will require the implementation of *Predict*

Given source-data *d_s_* and target-data *d_t_*, the obvious form of transfer learning is one that is defined by *Transfer*(*d_t_ Learn*(*d_s_*)). Here, a model is first constructed for the source data, and is then transferred to the target data. However, the literature suggests that better models for a dataset *d* may be obtained by *Transfer*(*d Learn*(*d_g_*)), where *d_g_* denotes a large generic dataset (like ImageNet). In the following section, we investigate the performance of model transfer using source-data (*d_s_*) from the brain and target-data (*d_t_*) from the lung, and *vice versa*.

## 3 Empirical Evaluation of Transferring Lesion Models

### 3.1 Aim(s)

Given a source-dataset *d_s_*, a target-dataset *d_t_*, and a (large) generic dataset *d_g_*, we distinguish between the following target models: (a) *Baseline*, denoting *Learn*(*d_t_*); (b) *Lesion-agnostic*, denoting *Transfer*(*d_t_|Learn*(*d_g_*)); and (c) *Lesion-augmented*, denoting *Transfer*(*d_t_|Transfer*(*d_s_|Learn*(*d_g_*))). For reasons of space, we will not consider *lesion-only* models (*Transfer*(*d_t_|Learn*(*d_s_*))).

Our aim is to compare the performance of models (a)–(c), using model transfer from brain-to-lung and lung-to-brain. We clarify our definition of performance in 4.

### 3.2 Materials

The experiments here use the following datasets:

**ImageNet data:** This constitutes *d_g_*, or the generic data used for model transfer. This dataset is a classical collection of images for visual recognition research [7].
**Lung-lesion data:** The LIDC-IDRI dataset contains lung CT scans with annotated lesions [6]. The malignancy level for each lesion is annotated in the range 1–5. The tumours with average malignancy values from 1 to 3 were considered as benign, and the rest were considered malignant.
**Brain-lesion data:** The brain tumour dataset [4] contains 3064 T1-weighted contrast-enhanced images from 233 patients with three kinds of brain tumours meningioma (708 slices), glioma (1426 slices) and pituitary tumour (930 slices) along with the corresponding lesion masks. We take meningioma and pituitary tumours as benign, and glioma tumours as malignant to form a binary classification problem.

### 4 Method

To help the model generalise better [2] we process the images by extract the lesions using the segmentation masks and normalise them. We then perform image augmentations such as random horizontal and vertical flips, shifts and rotations using the ImageDataGenerator class available in Keras [5].

Our experiments investigate the transfer of lesion models from brain-to-lung, and from lung-to-brain. We examine the effect of: (a) varying target training data size (with a fixed source data size); and (b) varying source data size (with a fixed target data size). For a given source (brain or lung), we adopt the following method:

Let *Te_t_* denote an independent sample of test instances for assessing the performance of the target model
**Fixed source data size:** For a given source data set *d_s_* and random samples *d_t_* of target data sizes in *{High, Medium, Low, V Low}*:

1. Construct *Base_t|s_* = *Learn*(*d_t_*)
2. Construct *LesAgnt|s* = *Transfer*(*d_t_|Learn*(*d_g_*))
3. Construct *LesAugt|s* = *Transfer*(*d_t_|Transfer*(*d_s_|Learn*(*d_g_*)))
4. Compare the performance of *Base_t|s_, LesAgnt|s*, and *LesAugt|s* on *Te_t_*
**Fixed target data size:** For a given target data set *d_t_* and random samples *d_s_* of source data sizes in *{High, Medium, Low, V Low}*:

1. Construct *Base_s|t_* = *Learn*(*d_t_*)
2. Construct *LesAgns|t* = *Transfer*(*d_t_|Learn*(*d_g_*))
3. Construct *LesAugs|t* = *Transfer*(*d_t_|Transfer*(*d_s_|Learn*(*d_g_*)))
4. Compare the performance of *Base_s|t_, LesAgns|t*, and *LesAugs|t* on *Te_t_*

The following details are relevant:

1. The overall sizes of the brain data and lung data are 3064 and 729 respectively. The sizes of independent test data is 200 images for both brain-to-lung and lung-to-brain transfer. In all cases *d_g_* refers to ImageNet.
2. The size of *d_s_* in Step 4 with brain-as-source is 3000 brain images, and *d_s_* with lung-as-source is 700 lung images. In all cases, we use the following target data sizes *d_t_*: 400 (High); 335 (Medium); 250 (Low); and 165 (VLow).
3. The size of *d_t_* in Step 4 with brain-as-target and lung-as-target is 400 images. For brain-as-source, the sizes of source data *d_s_* are: 3000 (High); 2000 (Medium); 1000 (Low); and 500 (VLow). For lung-as-source, the size of source data *d_s_* are: 700 (High); 500 (Medium); 300 (Low); and 100 (VLow).
4. In all our experiments, we use the model structure of DenseNet-201 [9] to construct the lesion classification models (source and target). The final structure and parameters of models are determined using the Adam optimiser [11] using a binary cross entropy loss function. Early stopping was used while monitoring the validation loss.
5. The training hyper-parameters for the algorithm and the models are as follows: the batch size is set to 64, the learning rate is 10^−4^.
6. All the experiments are conducted in Python environment in a machine with 64GB main memory, 16-core Intel processor and 8GB NVIDIA P4000 graphics processor.
7. We define the performance of a model as the pair (*R, P*) where *R* is an unbiased estimate of the recall of the model, and *P* is an unbiased estimate of the model’s precision.^4^ The performance of a pair of models will be compared lexicographically. That is, (*R*_1_*, P*_1_) is better than (*R*_2_*, P*_2_) if and only if: *R*_1_ *> R*_2_, or if *R*_1_ = *R*_2_ and *P*_1_ *> P*_2_.

## 5 Results

The principal findings of our experiments are these: (a) Target models obtained using lesion-augmented transfer perform better than those obtained using lesion-agnostic transfer in most of the cases as seen in Fig. 1; (b) As the lesion-augmented target data *d_t_* decreases, the benefit of lesion-augmented transfer over lesion-agnostic transfer increases (illustrated more clearly with *F*_2_ scores in Fig. 4, we note that the gap in the score is largest in the regime of very low target training data); and (c) As the source data *d_s_* decreases, the benefit of lesion-augmented transfer over lesion-agnostic transfer decreases as seen in Fig. 2. Surprisingly, we also note that as the source data decreases, sometimes the model ends up performing worse than the baseline models on the test set. We now highlight some additional aspects of our study: **Lower Variance of Lesion-Augmented Models:** We find that predictions using lesion-augmented models display a lesser variance than those from lesion-agnostic models. An example of this is shown in Fig. 3, showing the range of estimates of recall, precision obtained when performing a 5-fold cross-validation with *d_s_* = ‘High’, and *d_t_* = ‘High’. This suggests that these models depend less on a particular training instance. The same trend holds in lung-to-brain transfer, but is less pronounced. **Faster Convergence of Lesion-Augmented models:** We note that the gains of using a lesion-augmented model are twofold: (a) The loss starts from a much lower value, and (b) The lesion-augmented model converges in a fraction of the time that either of the other models takes to converge. An example of this is shown in Fig. 5 with the normalised binary cross-entropy loss. This behaviour occurs in lung-to-brain transfer as well, and we haven’t shown it here due to space limitations.

**Fig. 1.**
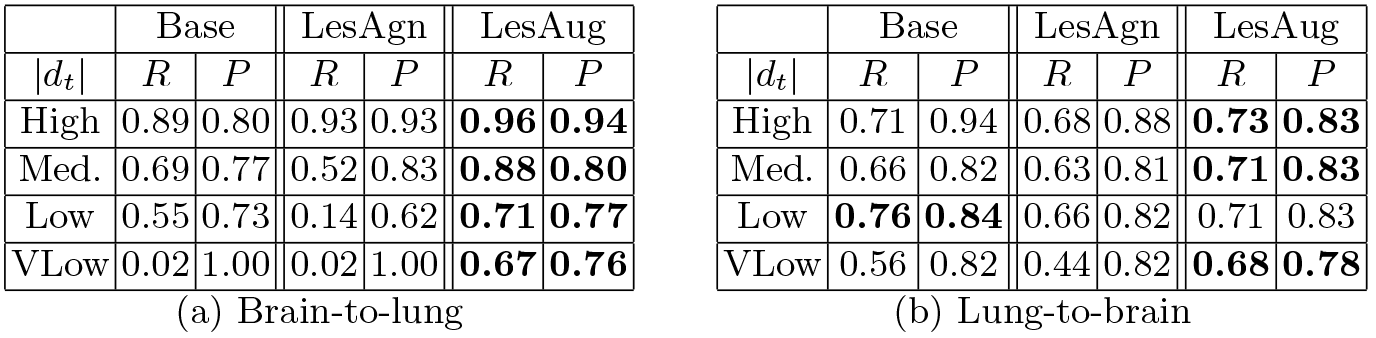
Recall and Precision scores keeping source data constant. The best performance pair (R, P) in each row is in bold.

**Fig. 2.**
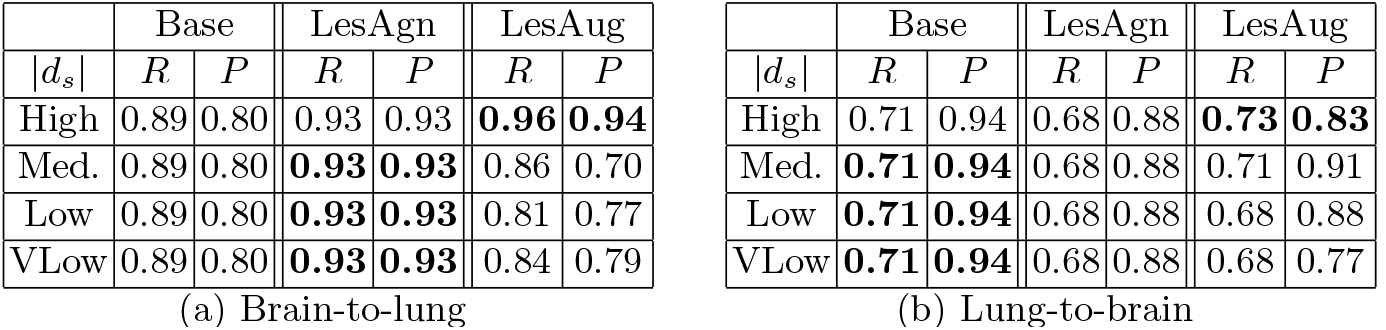
Recall and Precision scores keeping target data constant. ‘Base’ and ‘LesAgn’ models have no access to *dS*, therefore *R* and *P* scores are the same across |*d_s_*| values.

**Fig. 3.**
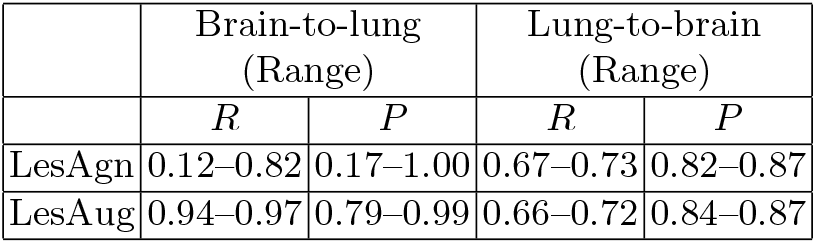
Ranges of recall(*R*) and precision(*P*) in a 5-fold cross-validation

**Fig. 4.**
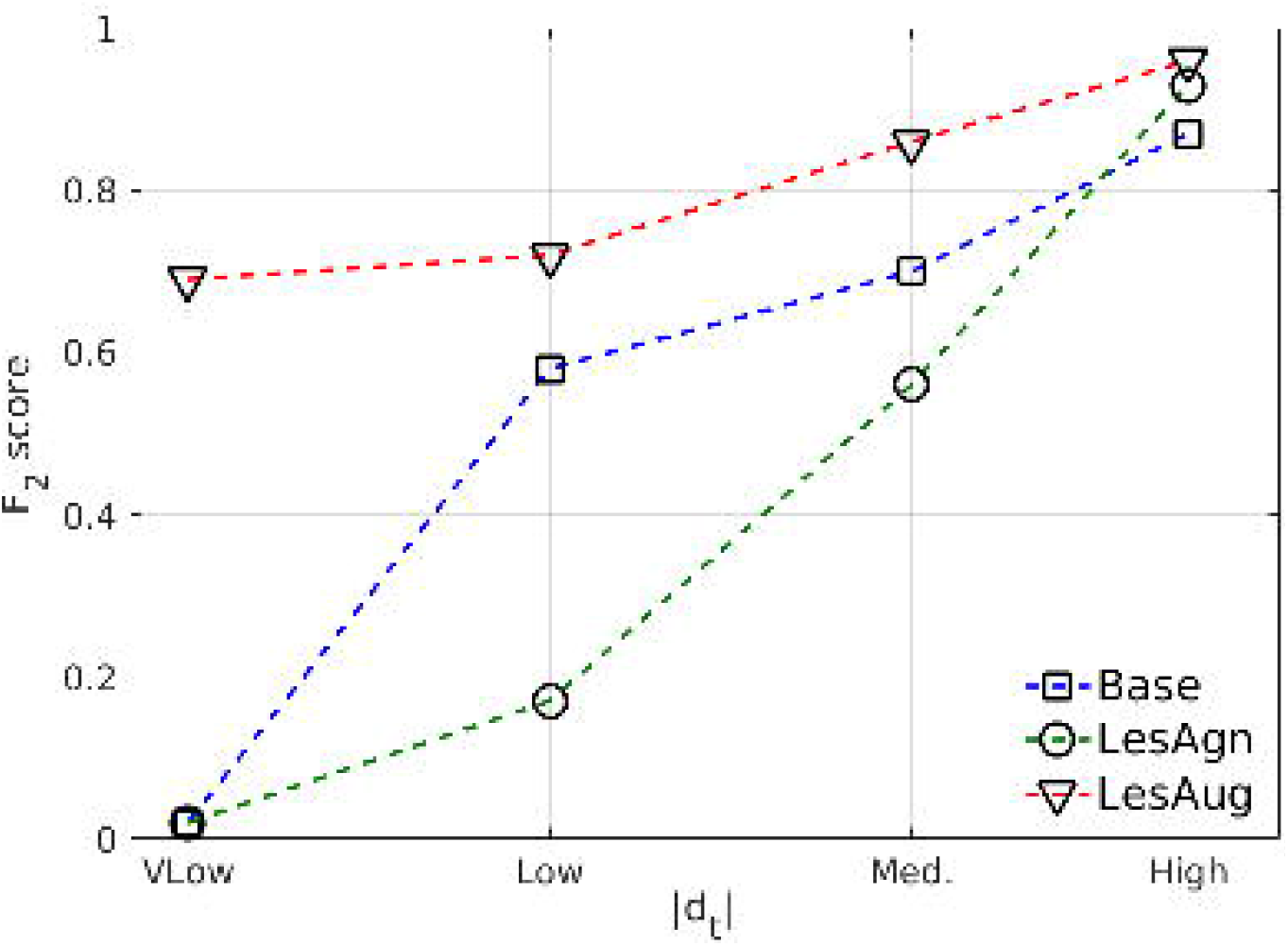
*F*2 score vs *d_s_* for brain-to-lung transfer, varying |*d_t_*|

**Fig. 5.**
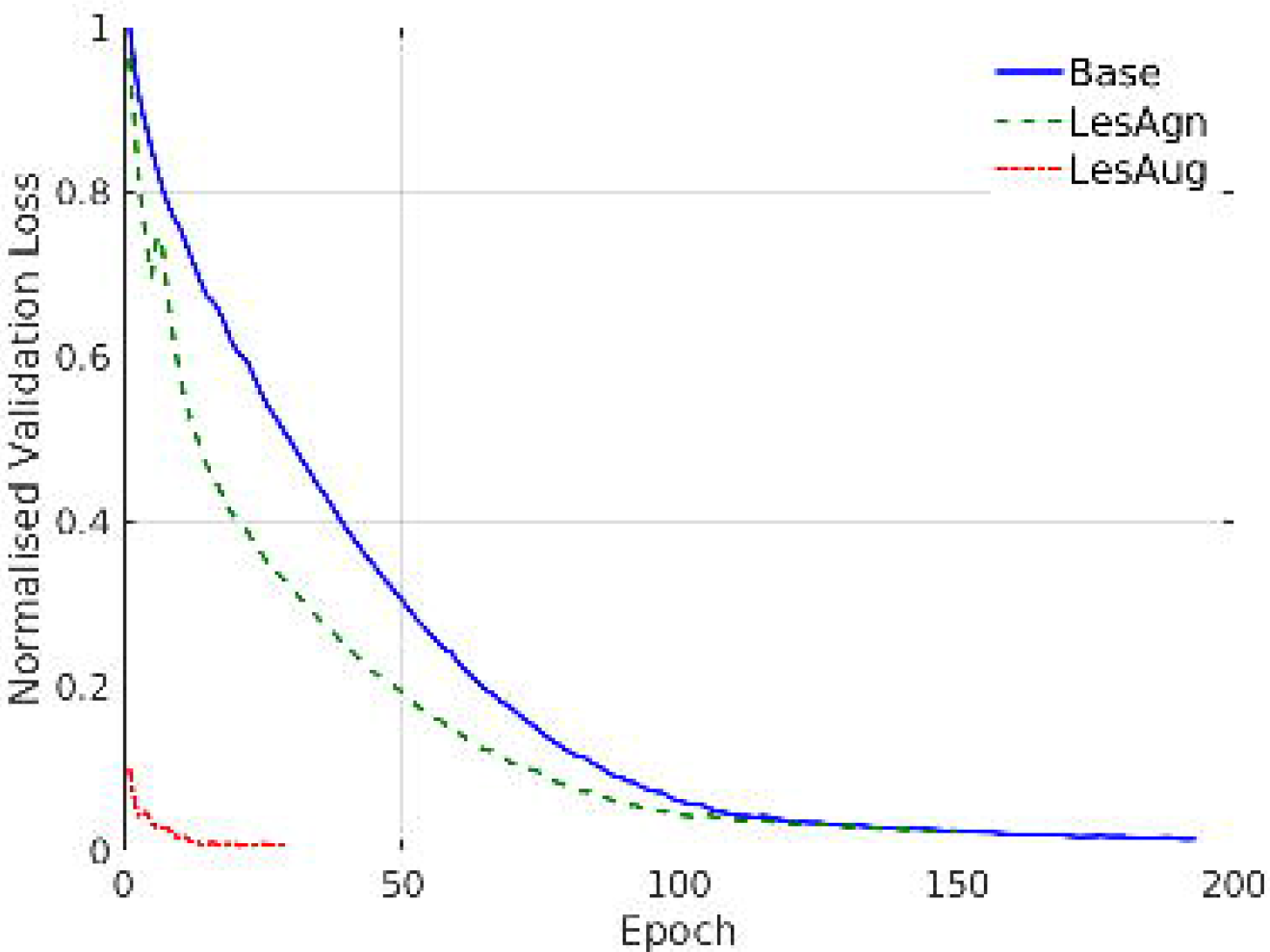
Loss vs epochs for brain-to-lung transfer, using a “VLow” value of *|*>d_t_*|*

## 6 Conclusion

Transfer learning, especially using deep neural networks, presents one way of dealing with the problems arising from the lack of sufficient data to build good models, that is common in problems involving medical images. This is due to reasons of cost, rarity of disease occurrence, difficulty of annotation, and so on. Although most routine demonstrations of transfer learning with deep networks have involved transfer from general datasets to specific ones, it would seem evident that transfer would be more effective if the source data were in some way related to the target. In this paper we have investigated this for transfer of lesion-models from one organ to another. Although the problem of lack of data persists for lesions for some organs, there are good biological reasons to believe that lesions from a different organ could be useful in constructing a model for the target organ. Our results here show how even small amounts of such lesion-specific source data can make a substantial difference to target models (the augmentation of a dataset of nearly 14m images, with at most 5000 lesion images for the source organ). Besides better predictive performance, we find that the augmentation results in target models that converge faster and have lower variance.

While our results are largely consistent for both brain-to-lung and lung-to-brain transfer, there is a difference in the gains resulting from the inclusion of lesion-specific data. Additional experiments suggest that this is not due to differences in the quantity or quality of data. There may be some underlying biological reasons for this difference, which needs to be investigated further. We think the experiments could also benefit from the use of data available in [13], which already has segmented lesions throughout the body. If annotations were available for even some small part of this data, they could prove more helpful than using a generic dataset like ImageNet. Additional experiments are also needed with other pairs of organs, to ensure that the observations here can be generalised: for the present, the results show that lesion-knowledge can be transferred usefully from the brain to the lung and *vice versa*.

## Data Availability

The data used in the paper is available online.

https://wiki.cancerimagingarchive.net/display/Public/LIDC-IDRI

https://figshare.com/articles/brain_tumor_dataset/1512427

## Acknowledgement

This work is supported by “The DataLab” agreement between BITS Pilani, K.K. Birla Goa Campus and TCS Research, India. AS is a Visiting Professorial Fellow at the School of CSE, UNSW Sydney.

## Footnotes

4 We assume that in lesion-identification, the positive class refers to malignant lesions and that false-negatives are costlier than false-positives. That is, recall is more important than precision.

